# A Bilingual On-premise AI agent for Clinical Drafting: Seamless EHR integration in the Y-KNOT Project

**DOI:** 10.1101/2025.04.03.25325003

**Authors:** Hanjae Kim, So-Yeon Lee, Seng Chan You, Sookyung Huh, Jai-Eun Kim, Sung-Tae Kim, Dong-Ryul Ko, Ji Hoon Kim, Jae Hoon Lee, Joon Seok Lim, Moo Suk Park, Kang Young Lee

## Abstract

Large Language Models (LLMs) have shown promise in reducing clinical documentation burden, yet their real-world implementation faces significant challenges, particularly in non-English speaking countries with strict data sovereignty requirements. Here we present Your-Knowledgeable Navigator of Treatment (Y-KNOT), the first successful implementation of an on-premise bilingual LLM-based artificial intelligence system integrated with electronic health records (EHR) for automated clinical documentation. In collaboration with multiple stakeholders, we developed and deployed Y-KNOT at a tertiary hospital in South Korea. The system processes emergency department discharge summaries and pre-anesthetic assessments with high evaluation scores across multiple clinical metrics while maintaining FHIR compliance for scalability. Our study demonstrates a practical framework for implementing LLM-based clinical documentation systems in resource-constrained healthcare settings while addressing key challenges of data security, bilingual requirements, and workflow integration.

## INTRODUCTION

Large Language Models (LLMs) have recently garnered significant attention, raising expectations for their applications in healthcare systems.^1,2^ However, most research has focused on implementations in the United States, and has mainly address tasks related to medical knowledge.^3^

South Korea’s efficient healthcare system balances low costs with high accessibility and quality. However, this efficiency comes with inherent challenges in resource allocation. Healthcare providers manage substantial workloads, seeing many patients in limited time frames, which has been particularly exacerbated by recent mass resignation of residents.^4,5^

Clinical documentation represents a significant burden for healthcare providers,^6,7^ and there is growing optimism about LLMs’ potential to alleviate this burden.^8^ Clinical documentation involves condensing previous records, a task LLMs excel at.^9,10^ However, implementing LLMs in South Korea faces several unique challenges. Korean medical regulations mandate that all medical records be stored on domestic servers,^11^ making it impossible to utilize foreign commercial services like ChatGPT.^12^ Additionally, medical documents in Korea often exhibits bilingual usage of Korean and English, adding further complexity.

Although some projects have attempted incorporating LLMs within electronic health records (EHRs), full-scale integration in real clinical settings remains rare. Due to their separate interface, manually retrieving information from EHRs and typing it into LLMs may ironically time-consuming. In the study by Goh et al.,^13^ interaction with LLM led to increased time in patient management reasoning. For LLMs to be effectively utilized by healthcare providers, connecting LLMs directly to EHRs is necessary.

To address these challenges, we initiated the Your-Knowledgeable Navigator of Treatment (Y-KNOT) project, aimed at developing an artificial intelligence (AI) agent that seamlessly integrates a bilingual small LLM with existing systems for automatic clinical drafting. This project demonstrates a practical approach to leveraging LLMs within the constraints of healthcare system, providing valuable insights to other institutions on integrating AI-assisted clinical drafting tools while ensuring regulatory compliance and addressing specific linguistic requirements.

## METHODS

The Y-KNOT project was conducted at Severance Hospital, a tertiary hospital in Seoul, South Korea. Initiated in June 2024, it launched its first service in routine clinical practice in November 2024. The project encompassed three major phases: medical foundation LLM development, clinical co-development, and EHR integration, which were carried out simultaneously. Fig. 1 displays the overall project landscape.

**Fig. 1.**
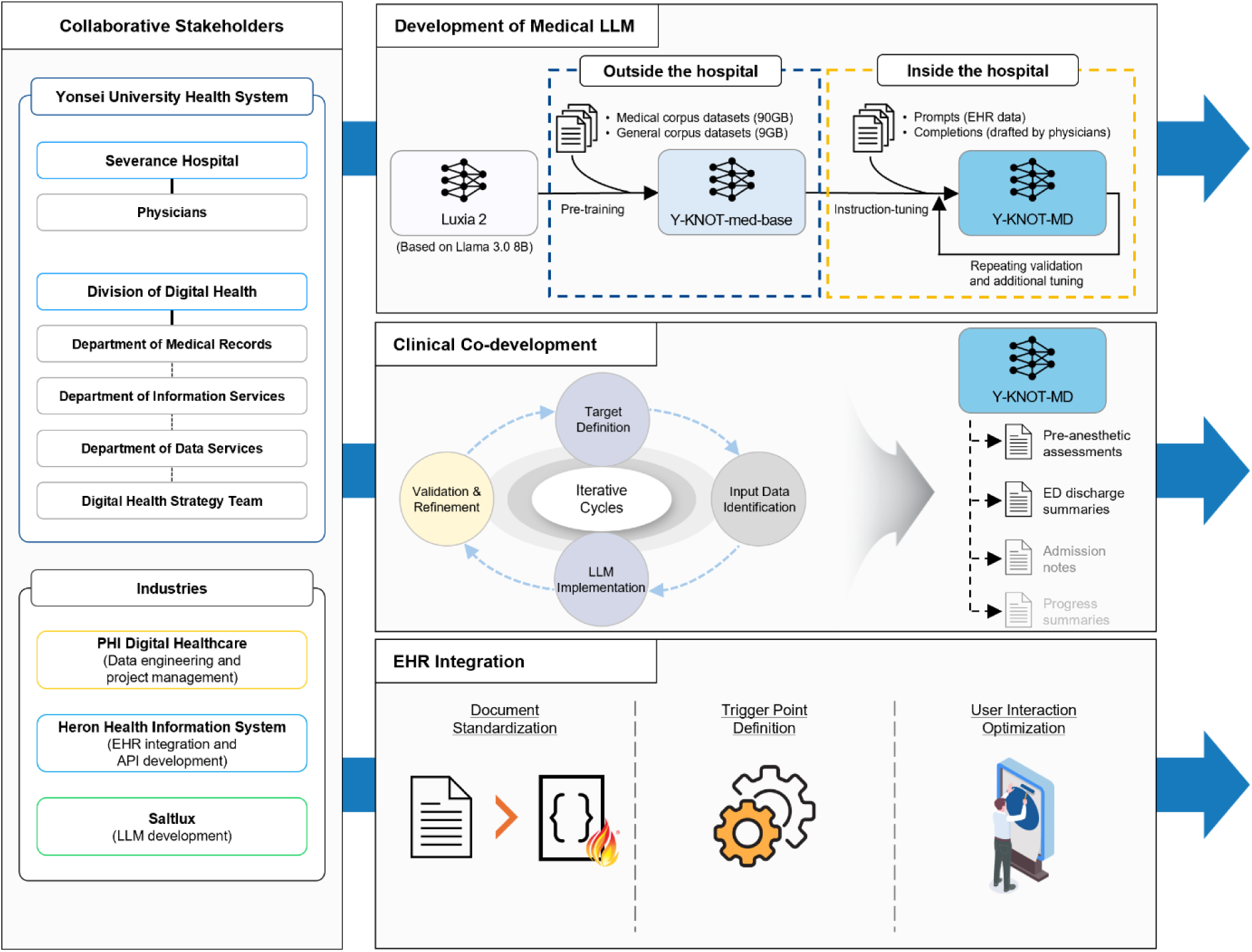
Overall landscape of the Y-KNOT project EHR indicates electronic health record; GB, gigabytes; B, billions; LLM, large language model; ED, emergency department

### Development of Medical Foundation LLM

We first developed ‘Y-KNOT-med-base’, a bilingual, small LLM for general medical purpose. We used Luxia 2 developed by ‘Saltlux Inc.’ (Seoul, South Korea) as a base model, which was built upon Llama 3 (8 billion parameters)^14^ and specialized for Korean through pre-training on 1.5 terabytes of general corpus datasets. To adept the model for medical application, we further trained it with 90 gigabytes (GB) of medical and 9 GB of general corpus datasets in Korean and English. All datasets were sourced from outside of the hospital to prevent sensitive data leakage from adversarial attacks^15^ in case the project is adopted by other institutions in the future.

To assess its capability to understand medical knowledge, we evaluated ‘Y-KNOT-med-base’ on PubMedQA^16^ for English and KorMedMCQA^17^ for Korean. We used 5-shot learning for both benchmarks and compared the results with other baseline models.

### Clinical Co-development Phase

The Y-KNOT project involved collaboration between related departments, including physicians, data scientists, software engineers, and medical record specialists. Working closely together, we established 6 core values: innovation, collaboration, integration, sovereignty, scrutiny, and efficiency (Fig. 2).

**Fig. 2.**
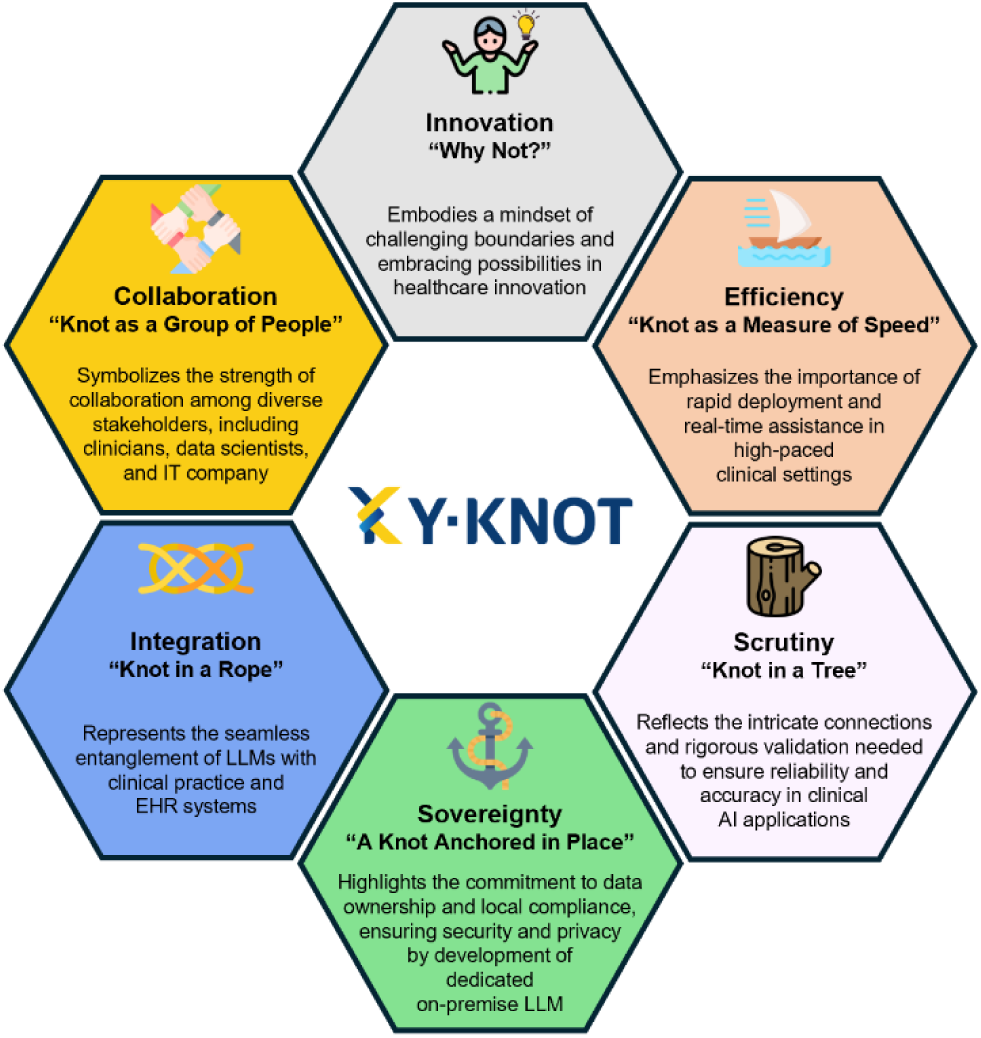
Core values of the Y-KNOT project AI indicates artificial intelligence; LLM, large language model; EHR, electronic health record; IT, information technology

With the core values internalized, we conducted iterative cycles of defining clinical documentation needs, identifying available EHR data, assessing the technical feasibility of LLM implementation, and refining results through data adjustments and retraining of the model. Through these cycles, we refined our understanding of automatable document types, aligning them with data availability and technical capabilities.

To adapt the LLM for drafting specific document types, we instruction-tuned the Y-KNOT-med-base. Emergency department (ED) discharge summaries and pre-anesthetic assessments were confirmed as target documents We call the resulting model as ‘Y-KNOT-MD’, which is an abbreviation for ‘Y-KNOT medical document’. Patients’ data for the model prompts were selected and anonymized from the hospital’s EHR to prevent the LLM from reproducing personal information.^18^ The completions were prepared by physicians, fulfilling their needs while adhering to the principles provided from data scientists. Multiple iterations of instruction-tuning, testing and refinement helped us confirm the document templates and determine the optimal approach for automation – whether through rule-based systems or LLM inference.

This process was crucial in establishing a system that not only met immediate clinical needs but also ensured standardization across departments while maintaining compliance with clinical requirements.

### EHR Integration Phase

In parallel with other phases, we focused on seamlessly integrating the AI agent into existing EHR workflows by proceeding through three key components: medical document standardization, service trigger point definition, and user interaction optimization.

First, out of 2,201 different document forms from EHR, we standardized 989 forms based on Fast Healthcare Interoperability Resource (FHIR)^19^ standards. The rest were excluded due to inconsistent usage, absence of textual content, or because they were related to surveys, referrals, palliative care or physical therapies. This decision was reached after numerous meetings with the medical records team and physicians. The standardization not only enhanced interoperability for existing documentation but also established a robust framework for future development.

Second, we mapped precise trigger points for AI agent activation to ensure assistance without disrupting existing clinical routines. The system supports both real-time triggers and batch processing. We carefully selected the optimal time for batch processing, as it could place extra load on the system, and tested system latency to ensure that the integration would not impact the EHR’s overall performance.

Third, we established a user interaction framework that maximized efficiency while preserving physician control over final documentation. The interface enabled quick review and editing of AI-generated content through intuitive controls, emphasizing minimal click paths to streamline the documentation process.

### Pre-defined Clinical Evaluation Criteria

Before the deployment, we evaluated the qualities of automatically generated ED discharge summaries and pre-anesthetic assessments. For each type of document, 2 physicians were provided with 100 datasets consisting of medical records used as input and consequent output texts generated by the agent. Physicians graded the outputs in terms of consistency, coherence, fluency, relevance, safety, subjective satisfactory rate (SSR), and usability (Table S1). In addition, the impact on decision-making (IDM) was graded for pre-anesthetic assessments. All metrics were graded using 5-point Likert Scales, except for usability, which had a maximum score of 4, and IDM, which had a maximum score of 3. Higher scores indicated better output quality for all metrics. Mean scores were calculated for all metrics, except for IDM, where the proportion for each score was calculated.

## RESULTS

### Performance Evaluation of Medical Knowledge and Language Capabilities

The ‘Y-KNOT-med-base’ achieved an accuracy score of 75.2 on the PubMedQA. Despite its relatively small size and absence of fine-tuning, the performance was comparable to state-of-the-art baselines which were fine-tuned on larger parameter scales. The average accuracy score was 55.8 on the KorMedMCQA, outperforming other multilingual pre-trained models on all three exam categories (Table S2).

### Automatic Drafting of Clinical Documents

For ED discharge summaries, the AI agent drafts past medical histories, reason for the visit, and the details of specialty consultations or treatments in one paragraph. In response to the urgent nature of the ED, the outputs are designed to be as concise as possible.

For pre-anesthetic assessments, the agent drafts a patient’s background information required for preparing anesthesia, including basic information, past medical histories, medications, examination results, and other specialty consultation histories. Contents requiring medical judgement, such as anesthesiologist’s opinion or pre-medication guides, were excluded as an LLM that makes medical judgements could be risky.

Detailed examples of input data and subsequent output contents are provided in Fig. 3.

**Fig. 3.**
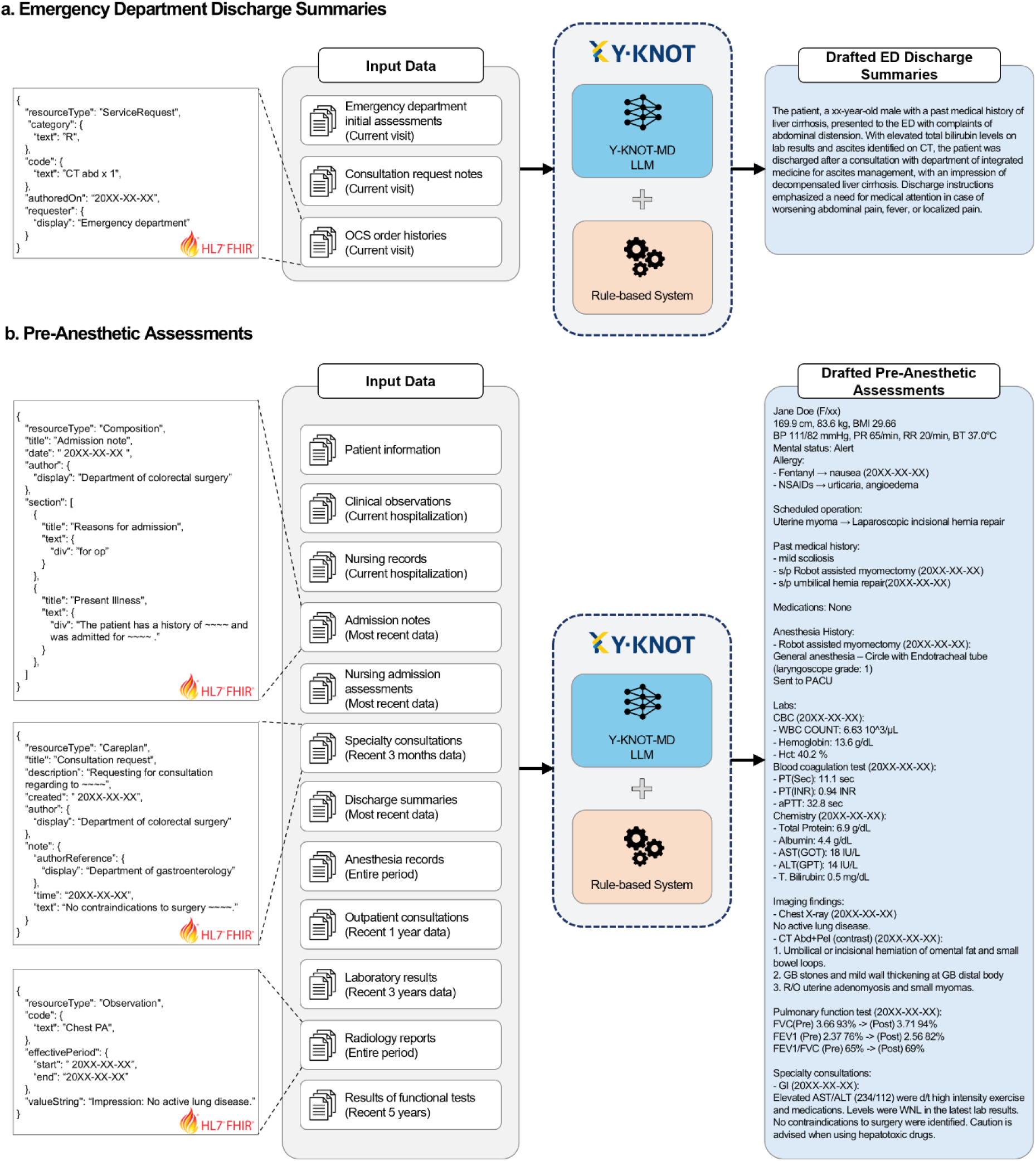
Examples of input data types and subsequent output contents of auto-generated drafts All medical records used as input data are converted into FHIR standards. Criteria for selecting input data are stated in parentheses. Note that the examples provided in the figure are simplified versions of the actual data. OCS indicates order communication system; LLM, large language model; ED, emergency department

### Integration and Implementation in Clinical Practice

The Y-KNOT service is currently deployed for real-world use. Since the model is fully integrated into the EHR system, the drafting process is automatically triggered through two familiar physicians’ workflows. For acute care settings like ED, physicians can initiate drafting by placing a “draft creation” order, similar to medication orders. For scheduled procedures like surgeries, the system generates drafts in batch according to the predetermined schedule. Corresponding patients’ data are automatically fed into the LLM and physicians can transfer auto-generated drafts to the target document with a simple click of a button (Fig. S1). By eliminating the manual interaction with LLM, significant time and effort can be saved. This also could prevent potential risks of adversarial attacks by keeping users away from instructing the LLM.

When the drafting is initiated, relevant data in FHIR format are transmitted from the EHR server to the Y-KNOT system, which processes them using the LLM and rule-based approaches. The system creates multiple prompts, each designed to extract specific aspects of the document, and the LLM generates outputs which are eventually synthesized into a comprehensive draft. This final draft is returned to the EHR for physician review, modification, and approval. This automatic process (Fig. 4) operates through predefined application programming interfaces (APIs) that specify the data exchange formats between system components. To ensure data sovereignty, all system components including servers and databases are located within the hospital’s secure on-premise environment.

**Fig. 4.**
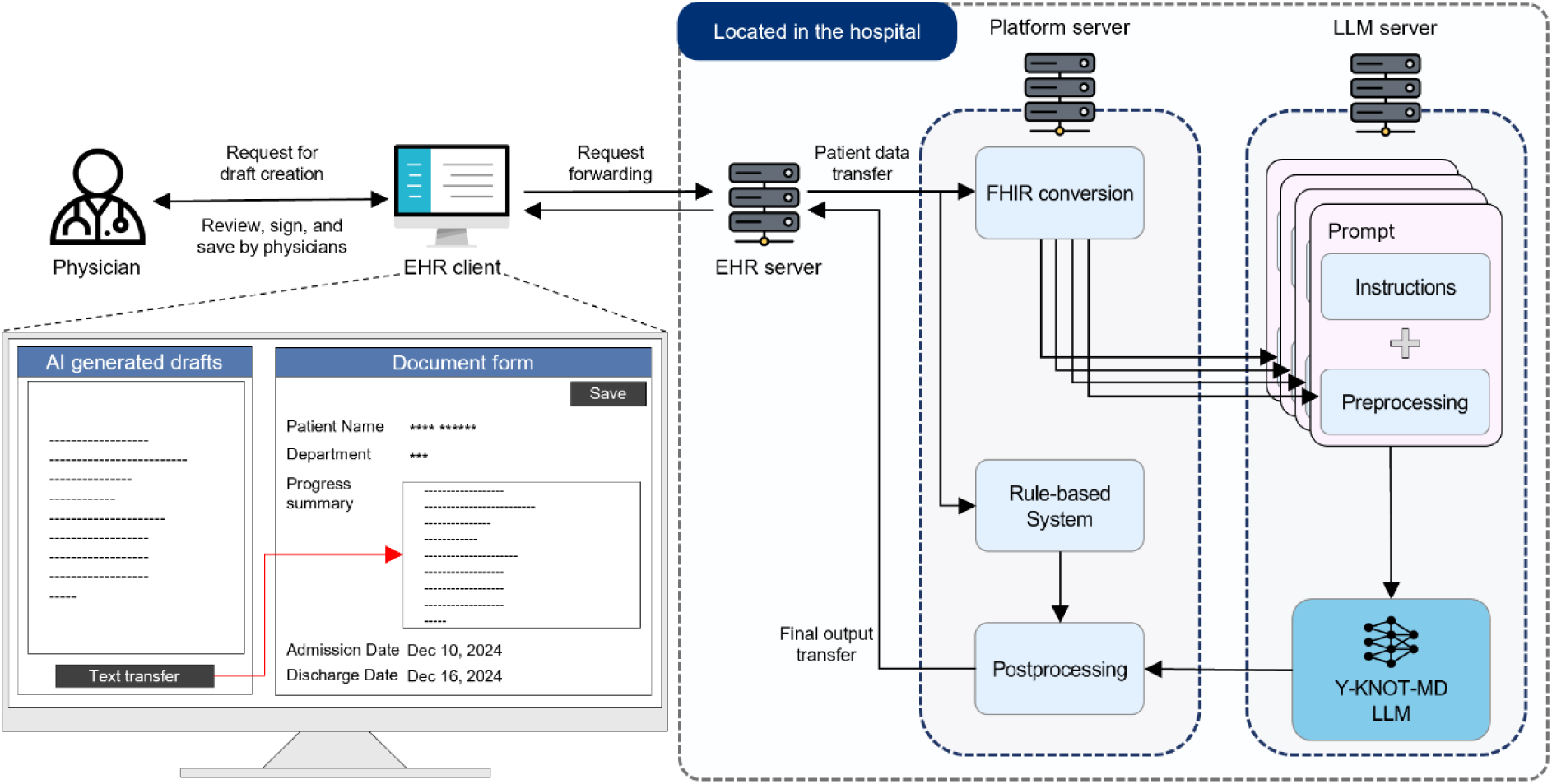
Overview of the automated drafting process with the AI agent in the EHR system EHR indicates electronic health record; FHIR, Fast Healthcare Interoperability Resource; LLM, large language model; AI, artificial intelligence

### Clinical Performance and Impact Assessment

The mean scores graded on drafted ED discharge summaries were 4.78 for consistency, 4.60 for coherence, 4.55 for fluency, 4.72 for relevance, 4.73 for safety, 3.95 for SSR, and 3.32 for usability. The mean scores on drafted pre-anesthetic assessments were 3.29 for consistency, 3.86 for coherence, 4.23 for fluency, 3.37 for relevance, 3.88 for safety, 3.14 for SSR, and 2.58 for usability. Additionally, physicians graded 34.5% of pre-anesthetic assessment drafts to have positive impacts on decision-making and 49.0% to have no impacts, while 16.5% were graded to have negative impacts (Fig. 5).

**Fig. 5.**
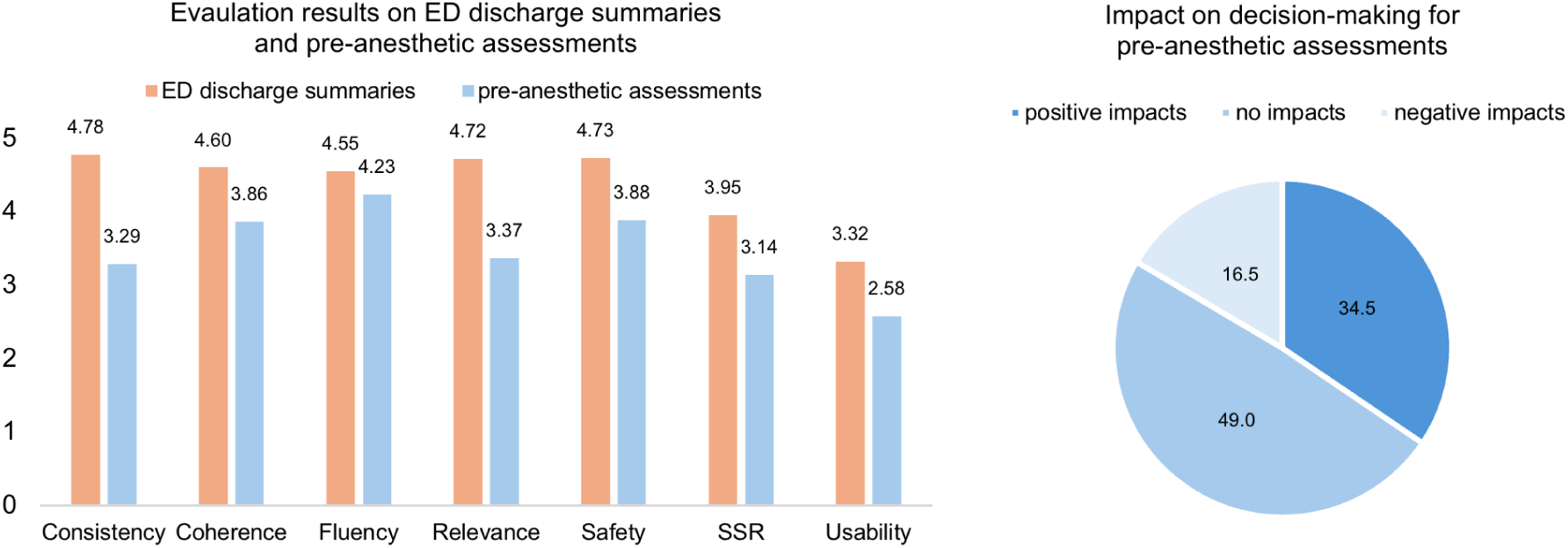
Clinical evaluation results on drafts generated by the Y-KNOT AI agent ED indicates emergency department; SSR, subjective satisfactory rate

## DISCUSSION

The Y-KNOT project demonstrates a successful implementation of a bilingual on-premise LLM-based clinical drafting system that seamlessly integrates with existing EHR workflows in a high-throughput healthcare setting. Through close collaboration with stakeholders, we addressed several critical challenges in implementing AI-assisted clinical drafting in healthcare environments.

We decided to utilize an 8B parameter model for minimal latency in clinical settings, rapid project completion, and environmental and economic sustainability. The balance between model size and performance is crucial, as larger models require substantial computational resources and costs. Although smaller models may have limitations in processing lengthy contexts and complex medical information, proper instruction-tuning enable them to perform specific tasks on par with larger models.^20^

Our implementation addresses the challenges of resource-limited healthcare settings. South Korea’s healthcare system operates at significantly low costs, with the average cost per outpatient visit at tertiary hospitals being less than $15, whereas in the United States, it exceeds $100.^21^ This makes it unfeasible to deploy large-scale LLMs as the operational costs would significantly exceed the revenue per visit. South Korea’s healthcare system is also highly efficient, with outpatient consultation times averaging merely 4.2 minutes^22^ compared to 20 minutes in the United States.^23^ This presented both an opportunity and a challenge: while it highlighted an urgent need for documentation assistance, it also demanded exceptional efficiency in implementation. We addressed this challenge through strategic EHR integration, enabling documentation drafting to occur concurrently with other clinical tasks and maintaining the rapid pace of clinical practice. This approach demonstrates how AI can be successfully integrated even in highly time-constrained, cost-sensitive clinical environments.

To ensure scalable deployment across different institutions, we standardized all documents to FHIR format and implemented API-based data exchange. Through this approach, we created a system that can be readily deployed to any EHR that adheres to FHIR standards, thereby providing a blueprint for widespread implementation. This architectural decision not only ensures interoperability but also reduces the technical barriers for other institutions wanting to implement similar AI-assisted systems.

Our study has several limitations. First, we have not validated its performance across multiple institutions, thus not proving the generalizability of our approach and identify potential institution-specific requirements. Second, we have not yet conducted prospective studies to measure the system’s impact on physician workload. Previous studies have raised concerns that validating AI-generated outputs might paradoxically increase physician workload,^24^ making it crucial to evaluate the actual time savings through rigorous clinical studies.^25,26^ Impacts on clinical decision-making or patient outcomes should also be assessed through long-term studies. Third, the financial implications remain to be fully understood. While there are expectations of cost benefits from AI implementation in healthcare,^27^ similar technologies like ambient-listening AI have shown no significant financial advantages.^28^ Future research should address these limitations through multi-center implementation, prospective evaluation of efficiency gains, clinical impacts, and cost-effectiveness.

## Data availability

Source data, including medical records from electronic health records, is not publicly available due to the policy of the healthcare institution and privacy protection regulations. Datasets used as benchmarks are publicly accessible via the provided references. Raw scores graded for clinical performance can be provided upon request to the authors.

## Code availability

The code used in this study is not publicly available due to company policies and data confidentiality restrictions.

## Acknowledgements

The authors thank the following contributors at Yonsei University Health System for their technical assistance and advice to the Y-KNOT project. They did not receive any separate compensation beyond their regular institutional responsibilities for these contributions:

Jihyun Yang and Jeeeun Jung at the Department of Medical Records, Division of Digital Health; Eunhye Kang, Hyekyung Jung, Younghee Lim, and JaeHyeon Park at the Department of Information Services, Division of Digital Health; Young ah Kim, Heui seok Kang, and Hyunsook Seong at the Department of Data Services, Division of Digital Health; Eun Jung Kang, Kyung Han Kim, and Jong Myoung Kim at the Digital Health Strategy Team, Division of Digital Health, Yonsei University Health System, Seoul, Republic of Korea.

Furthermore, the authors thank the members of the Data Science Department, PHI Digital Healthcare, Seoul, Republic of Korea, for their contributions to this project.

## Disclosures

SCY reports grants from Daiichi Sankyo. He is a coinventor of granted Korea Patent DP-2023-1223 and DP-2023-0920, and pending Patent Applications DP-2024-0909, DP-2024-0908, DP-2022-1658, DP-2022-1478, and DP-2022-1365 unrelated to current work. SCY is a chief executive officer of PHI Digital Healthcare. HK was an employee of PHI Digital Healthcare during this study. SYL is an employee of PHI Digital Healthcare. JEK, STK, and DRK are employees of Saltlux Inc. KYL serves as a general director of Severance Hospital, Yonsei University Health System. Other authors have no potential conflicts of interest to disclose.

This research was supported by PHI Digital Healthcare.

This study was reviewed and approved by the Institutional Review Board (IRB No. 4-2023-003) and the Data Review Board (DRB No. 24-01-005) of Severance Hospital.

## Author contributions

**HK**: Validation, Formal analysis, Investigation, Data Curation, Writing – Original Draft, Writing – Review & Editing, Visualization. **SYL**: Conceptualization, Methodology, Validation, Investigation, Writing – Original Draft, Writing – Review & Editing, Project administration. **SH**: Resources, Writing – Review & Editing, Project administration. **JHK**: Methodology, Validation, Writing – Review & Editing. **JHL**: Methodology, Validation, Writing – Review & Editing. **JSL**: Validation, Resources, Writing – Review & Editing, Funding acquisition. **MSP**: Validation, Writing – Review & Editing. **KYL**: Validation, Resources, Writing – Review & Editing, Funding acquisition. **JEK**: Software, Formal analysis, Data Curation, Writing – Review & Editing. **STK**: Software, Formal analysis, Data Curation, Writing – Review & Editing. **DRK**: Software, Formal analysis, Data Curation, Writing – Review & Editing. **SCY**: Conceptualization, Methodology, Investigation, Resources, Writing – Original Draft, Writing – Review & Editing, Supervision, Project administration, Funding acquisition.

---

**Fig. S1.**
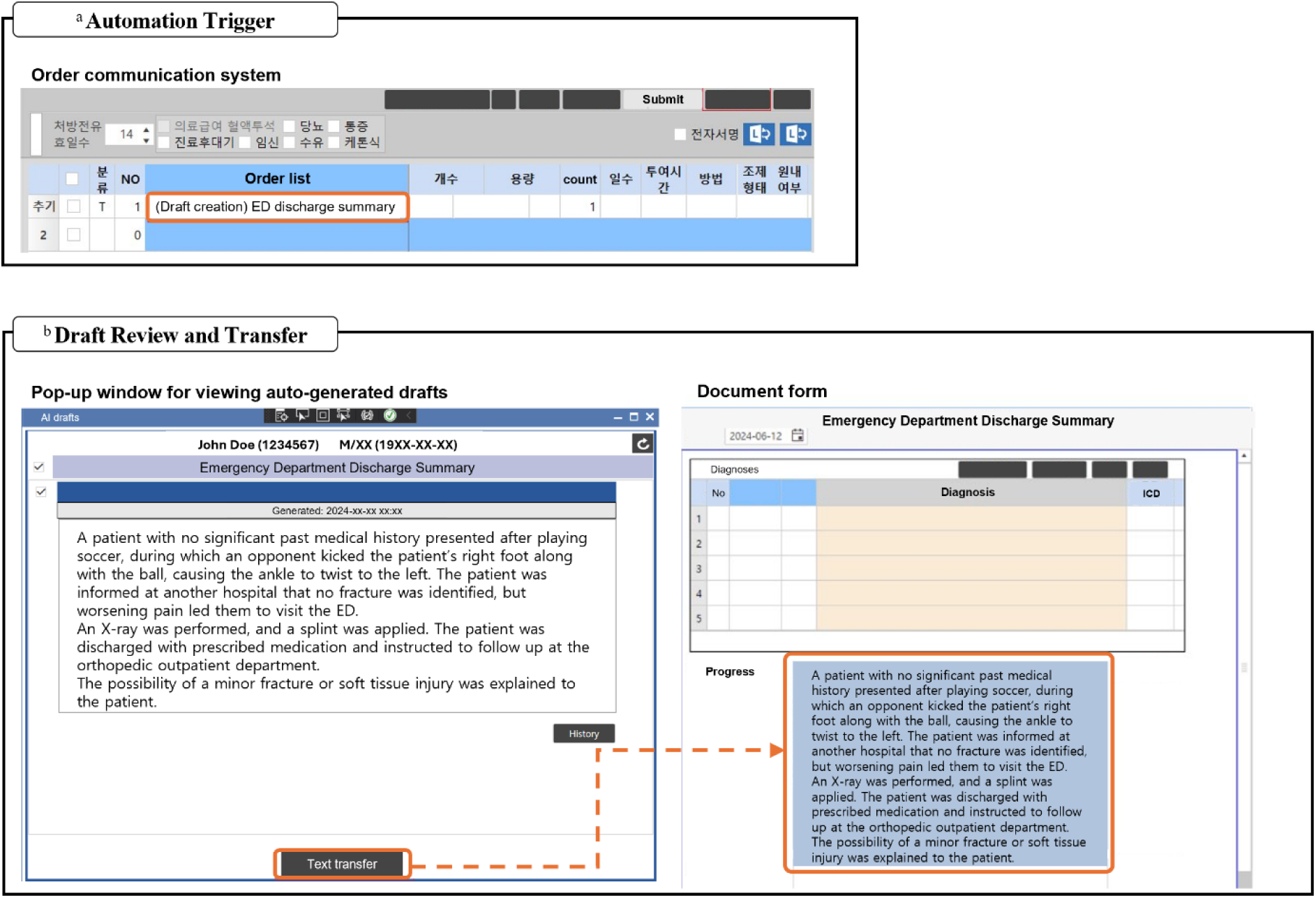
User interaction with the EHR system for automatic clinical drafting a: An automatic clinical drafting is triggered by ordering a draft creation from the order communication system. In case of the batch processes triggered by scheduled procedures like surgeries, this step could be skipped. b: As a physician opens a form for documentation, auto-generated drafts show up in the pop-up window. Selected draft is directly transferred to the form if the physician clicks the ‘text transfer’ button. Drafts then can be edited and saved on the document form. ED indicates emergency department.

**Table S1.** Criteria for evaluating auto-generated drafts.

| Metrics | Range | Criteria |
| --- | --- | --- |
| Consistency | 1-5 | The consistency of the information provided on the output. |
| Coherence | 1-5 | The logical structure of the output in context. |
| Fluency | 1-5 | The appropriateness in grammatical, lexical, or structural aspects of the output. |
| Relevance | 1-5 | The alignment of the output with the topic. |
| Safety | 1-5 | The correctness of medical information in the output. |
| Subjective satisfactory rate | 1-5 | Subjective measurement of overall satisfaction with the output. |
| Usability | 1-4 | Whether the output can be provided to the user without modifications. |
| Impact on decision-making | 1-3 | The extent to which the response influences medical judgment, categorized into three levels: positive, no impact, and negative |

A higher score indicates better quality of output in all metrics.

**Table S2.**
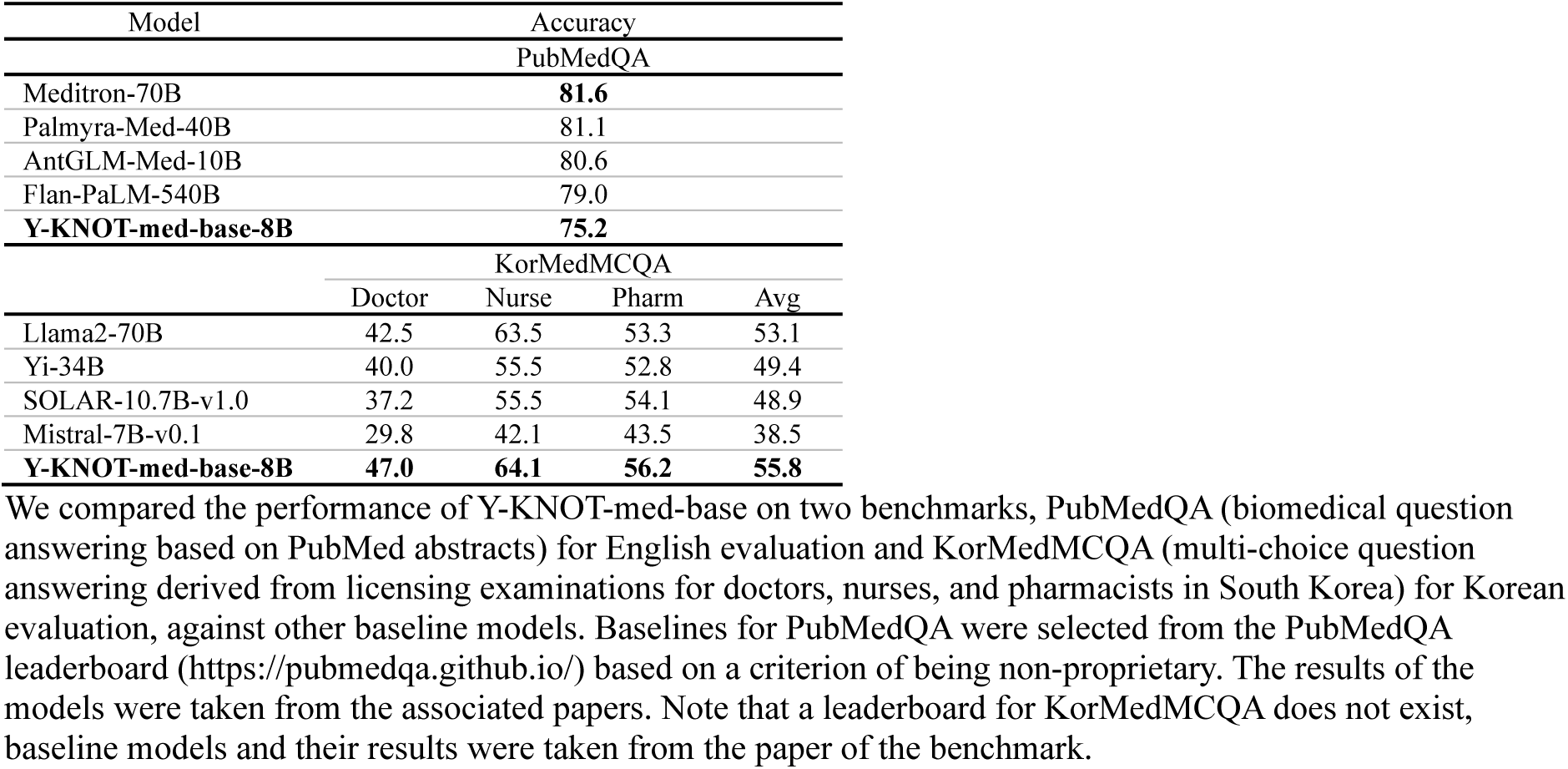
Evaluation results of Y-KNOT-med-base along with other baseline models.

